# Cortical lesions uniquely predict motor disability accrual and form rarely in the absence of new white matter lesions in multiple sclerosis

**DOI:** 10.1101/2023.09.22.23295974

**Authors:** Erin S Beck, W Andrew Mullins, Jonadab dos Santos Silva, Stefano Filippini, Prasanna Parvathaneni, Josefina Maranzano, Mark Morrison, Daniel J Suto, Corinne Donnay, Henry Dieckhaus, Nicholas J Luciano, Kanika Sharma, María Ines Gaitán, Jiaen Liu, Jacco A de Zwart, Peter van Gelderen, Irene Cortese, Sridar Narayanan, Jeff H Duyn, Govind Nair, Pascal Sati, Daniel S Reich

## Abstract

**Background and objectives:** Cortical lesions (CL) are common in multiple sclerosis (MS) and associate with disability and progressive disease. We asked whether CL continue to form in people with stable white matter lesions (WML) and whether the association of CL with worsening disability relates to pre-existing or new CL.

**Methods:** A cohort of adults with MS were evaluated annually with 7 tesla (T) brain magnetic resonance imaging (MRI) and 3T brain and spine MRI for 2 years, and clinical assessments for 3 years. CL were identified on 7T images at each timepoint. WML and brain tissue segmentation were performed using 3T images at baseline and year 2.

**Results:** 59 adults with MS had ≥1 7T follow-up visit (mean follow-up time 2±0.5 years). 9 had “active” relapsing-remitting MS (RRMS), defined as new WML in the year prior to enrollment. Of the remaining 50, 33 had “stable” RRMS, 14 secondary progressive MS (SPMS), and 3 primary progressive MS. 16 total new CL formed in the active RRMS group (median 1, range 0-10), 7 in the stable RRMS group (median 0, range 0-5), and 4 in the progressive MS group (median 0, range 0-1) (p=0.006, stable RR vs PMS p=0.88). New CL were not associated with greater change in any individual disability measure or in a composite measure of disability worsening (worsening Expanded Disability Status Scale or 9-hole peg test or 25-foot timed walk). Baseline CL volume was higher in people with worsening disability (median 1010μl, range 13-9888 vs median 267μl, range 0-3539, p=0.001, adjusted for age and sex) and in individuals with RRMS who subsequently transitioned to SPMS (median 2183μl, range 270-9888 vs median 321μl, range 0-6392 in those who remained RRMS, p=0.01, adjusted for age and sex). Baseline WML volume was not associated with worsening disability or transition from RRMS to SPMS.

**Discussion:** CL formation is rare in people with stable WML, even in those with worsening disability. CL but not WML burden predicts future worsening of disability, suggesting that the relationship between CL and disability progression is related to long-term effects of lesions that form in the earlier stages of disease, rather than to ongoing lesion formation.

## Introduction

In progressive multiple sclerosis (MS), there is gradual accrual of disability over time, often in the absence of new white matter or spinal cord lesions. This accrual of disability may be related to long-term effects of focal demyelination that occurs early in disease, persistent inflammation either diffusely or within pre-existing lesions, and/or ongoing focal demyelination that is not detected on standard MRI, including in the cerebral cortex.^1, 2^

Cortical MS lesions are associated with disability and progression, possibly to a greater extent than white matter lesions.^3–6^ Cortical lesions, and in particular subpial cortical lesions, which involve the superficial cortical layers, have been hypothesized to form via a partially distinct mechanism from other MS lesions that is related to inflammation in the overlying meninges.^7–9^ Despite their acknowledged prevalence on histopathologic studies, where a median of 14% of the cortex is demyelinated,^10–12^ cortical lesions have until recently been difficult to visualize on in vivo MRI. This has limited our understanding of when they develop and how they contribute to disability. Specifically, it is unclear whether the association between cortical lesions and progression is due to a persistent and long-term effect of cortical lesions that form early in the disease course or to ongoing cortical lesion formation in later stages of the disease. Intriguingly, recent studies have indicated that cortical lesion formation may be higher in progressive MS than in relapsing remitting MS (RRMS),^13, 14^ in contrast to white matter lesion formation, the vast majority of which occurs in the relapsing phase of the disease.

In this study, we followed three groups longitudinally — people in whom new white matter lesions had formed in the year prior to their enrollment (active RRMS); people with RRMS with stable white matter lesions in the year prior to enrollment (stable RRMS); and people with progressive MS with stable white matter lesions in the year prior to enrollment — with the goal of determining whether cortical lesions continue to form in people with stable white matter lesions and whether the association of cortical lesions with worsening disability is related to pre-existing or new cortical lesions. We found, surprisingly, that cortical lesion formation is rare in people with stable white matter lesions, even in those with worsening disability. Importantly, cortical but not white matter lesion burden predicts future worsening of disability, suggesting that the relationship between cortical lesions and disability progression is related to long-term effects of lesions that formed in the earlier stages of disease, rather than to ongoing lesion formation.

## Methods

### Clinical cohort

Participants enrolled in an institutional review board-approved MS natural history study at the National Institutes of Health provided written, informed consent. We built a prospective subcohort of MS patients who were ≥18 years old and without 7T MRI contraindication. Between May 2017 and November 2018, we prospectively enrolled individuals with stable white matter and spinal cord lesions by 3T MRI in the year prior to enrollment (stable MS), including 36 with RRMS and 19 with progressive MS (15 secondary progressive (SPMS) and four primary progressive (PPMS)). For comparison, we also enrolled nine individuals with RRMS who had at least one new or contrast-enhancing white matter lesion in the year prior to enrollment (active MS). As we did not have any data on rate of cortical lesion accumulation using our more sensitive 7T techniques at the time the study was initiated, sample size was driven by recruitment feasibility in our center, and we prioritized recruiting participants with radiographically stable MS.

Participants were evaluated annually for two years, including a clinical history and physical exam, expanded disability status scale (EDSS), paced auditory symbol addition test (PASAT), symbol digit modality test (SDMT, paper based), 25-foot timed walk (25TW), and 9-hole peg test (9HPT), a 7T brain MRI, and a 3T brain and spinal cord MRI. If obtained more than one day apart, the 25TW and 9HPT were performed at both 3T and 7T visits, and results were averaged. After the first two years, participants were followed annually with the same clinical testing for up to two additional years. Clinical relapses were determined based on chart review by an MS specialist.

### MRI acquisition

All 3T scans were acquired on a single Magnetom Skyra scanner (Siemens, Erlangen, Germany), equipped with a 32-channel head coil. The first 17 baseline scans were acquired prior to a software upgrade (D13 to E11). 3T brain scans included axial 2D proton density (PD)/T_2_w, sagittal 3D magnetization prepared 2 rapid gradient echo (MP2RAGE), and sagittal 3D T_2_w fluid attenuated inversion recovery (FLAIR) before and after administration of gadobutrol (unless contraindicated or refused by the participant). Several upgrades to the MP2RAGE version on the 3T scanner occurred in the first year of the study, after which there were no further changes in MP2RAGE version. Spinal cord scans included sagittal 2D short-tau inversion recovery (STIR) and sagittal 3D T_1_w gradient-recalled echo (GRE), sagittal 3D MP2RAGE of the cervical spine, and axial 3D MP2RAGE of the thoracic spine.

All 7T brain scans were performed on a 7T whole-body research system (Siemens) equipped with a single-channel transmit, 32-channel phased array receive head coil. A software upgrade (B17 to E12) was performed prior to the final five year-2 scans. 7T scans included axial 3D MP2RAGE (0.5mm isometric, acquired four times per scan session), sagittal 3D MP2RAGE (0.7mm isometric), and sagittal 3D segmented T_2_*w echo-planar imaging (EPI) (0.5mm isometric, acquired in two partially overlapping volumes for full brain coverage). At the baseline and year-1 timepoints, axial 2D T_2_*w multi-echo GRE was also acquired. At the year-2 timepoint, for 30 scans (all scans acquired after April 2020), an axial 3D T_2_*w multi-echo GRE with navigator-based B_0_ and motion correction was acquired in place of the 2D T_2_*w multi-echo GRE, as we found that this sequence improved image quality.^15, 16^ Both types of T_2_*w GRE scans were 0.5mm isometric and acquired in three partially overlapping volumes for near full supratentorial brain coverage. MP2RAGE data were processed into uniform denoised images (hereafter T_1_w MP2RAGE) and T_1_ maps using manufacturer-provided software. 7T images did not fully cover the inferior temporal and occipital lobes.

Median interval between 3T and 7T MRI was 27 days (IQR 61, range 1–294). Sequence parameters are listed in Supplementary Table 1.

### Image processing

The 7T 0.5mm T_1_w and T_1_ map MP2RAGE repetitions were coregistered and median T1w and T1 map images were generated, as described previously.^17^ The 7T T_2_*w GRE magnitude images were averaged across echo times, and within each timepoint, the averaged GRE images and the T_2_*w EPI images were linearly registered (FMRIB’s Linear Image Registration Tool, FLIRT^18, 19^) to the 7T T_1_w MP2RAGE median images. For the year-1 and year-2 timepoints, 7T T_1_ MP2RAGE median images were then registered to the baseline T_1_w MP2RAGE (Analysis of Functional NeuroImages, AFNI, 3DAllineate^20, 21^), and the transformation matrix was applied to the T_2_*w EPI and GRE magnitude images. Subtraction images were generated between registered T_1_ map images at each timepoint.

Baseline 3T images were processed and registered as described previously.^22^ For follow-up timepoints, FLAIR images were registered to T_1_w MP2RAGE (FLIRT), followed by registration of all FLAIR and T_1_w MP2RAGE to the baseline T_1_w MP2RAGE (AFNI, 3DAllineate).

### Longitudinal cortical and white matter lesion segmentation

Cortical lesions were manually segmented on baseline images using median 7T T_1_w and T_1_ map MP2RAGE, T_2_*w GRE, and T_2_*w EPI images by two independent raters as previously described.^23^ Lesions were categorized as leukocortical (type 1, involving cortex and white matter), intracortical (type 2, confined to the cortex and not touching the pial surface of the brain), and subpial (type 3, involving the cortex exclusively and touching the pial surface, and type 4, involving cortex and the white matter and touching the pial surface).^10^ Cortical lesions were hypointense on T_1_w MP2RAGE images and/or hyperintense on T_2_*w images and were seen on at least two consecutive axial slices. Cortical lesion volumes and median T_1_ within lesions, including the white matter portion of leukocortical lesions, were calculated.

For each subsequent timepoint, MP2RAGE and T_2_*w images were evaluated by two independent raters for the presence of new cortical lesions compared to the baseline scan. Cortical lesions that were visible at year 1 or 2 and whose presence at earlier timepoints was potentially obscured by motion were not segmented. In addition, the mask for each cortical lesion present at baseline was evaluated and adjusted at each timepoint both for imperfect registration as well as for changes in lesion size.

White matter lesions were segmented on baseline 3T images as previously described.^23^ Baseline white matter lesion masks were adjusted using registered year-2 T_1_w MP2RAGE and FLAIR images, and new lesions were identified and segmented. Longitudinal 7T MP2RAGE images were also assessed for new white matter lesions. Occasionally, small, new white matter lesions were identified only on 7T images but were seen in retrospect on 3T images. Once identified on 7T images, these new lesions were segmented on the corresponding 3T images.

### Spinal cord lesion assessment

Spinal cord lesions were identified by two independent raters followed by consensus review using sagittal STIR and MP2RAGE images of the cervical spinal cord and sagittal STIR and axial MP2RAGE images of the thoracic spinal cord at baseline and year 2.

### Brain volume and brain atrophy measurement

Brain volume measurements were obtained from 0.7mm T1w MP2RAGE images at all available 7T timepoints using Pseudo-Label Assisted “no-new-Net” version of U-Net algorithm (PLAn).^24^ PLAn is a transfer-learning method that uses pseudo-labels obtained from C-DEF segmentation of 3T images^25^ which were then fine tuned on MP2RAGE images from 7T in a separate cohort of patients. Retraining of PLAn was not required herein as the segmentation model trained on images from the scanner already existed. PLAn segmentations were assessed for quality and adjusted manually as needed. 0.7mm MP2RAGE was not available for all participants at all timepoints. For 47 participants, 0.7mm MP2RAGE was acquired at baseline and at least one follow-up timepoint. 10 scans from 5 individuals were discarded due to poor segmentation, leaving a total of 42 individuals with multiple timepoints available to analyze. Lesions identified by PLAn were filled and cortex, white matter, and deep gray matter volumes were extracted. Because of inconsistent infratentorial coverage in our 7T images, the brainstem and cerebellum were not included in the brain volume measurements. Baseline brain volumes were normalized using the MNI152 intracranial volume (https://nist.mni.mcgill.ca/icbm-152lin/).

To determine the rate of atrophy, annual atrophy rates were calculated from raw brain volumes at each timepoint (baseline to year 1 and year 1 to year 2), which were then averaged across timepoints for each individual.

### Statistical analyses

Comparisons between stable RRMS and PMS and between people with stable vs new lesions were performed using t-tests, Fisher’s exact test, and Mann-Whitney tests as appropriate.

Comparisons between active RRMS, stable RRMS, and PMS were performed using Kruskal-Wallis tests and one-way ANOVA as appropriate.

To investigate the relationship between baseline MRI measures and subsequent disability change and brain atrophy, we first performed Box-Cox transformation of changes in disability measures and lesion counts and volumes, and then calculated partial correlation coefficients, adjusting for age and sex, with 5000 nonparametric bootstrapping iterations to increase analysis robustness.^26, 27^

Comparisons between people with progression of disability, defined as an increase in EDSS of ≥1 for baseline EDSS <6 or ≥0.5 for baseline EDSS ≥6 or 20% increase in 25TW or 20% increase in 9HPT, vs stable disability and between people who transitioned from RRMS to SPMS vs those who remained RRMS were performed using a multivariable generalized linear model adjusted for age and sex. Box-Cox transformation of lesion counts and volumes was performed prior to running these models.

Adjustment for multiple comparisons was performed using the Benjamini and Hochberg procedure.^28^ Statistical analyses were performed on IBM SPSS Statistics, version 26 (IBM Corp., Armonk, NY, USA).

## Results

### Cortical lesion formation is rare in people with stable white matter lesions and is not associated with worsening disability

64 individuals with MS, including 9 active RRMS, 36 stable RRMS, and 19 progressive MS (15 SPMS, 4 PPMS), underwent baseline clinical evaluation, 3T brain and spinal cord MRI, and 7T brain MRI. The results of the baseline analyses have been previously described.^23^ 59/64 participants (92%) returned for at least one 7T follow-up visit (9 active RRMS, 33 stable RRMS, and 17 progressive MS [14 SPMS, 3 PPMS]) (Table 1). Mean ± standard deviation (SD) total 7T follow-up was 2 ± 0.5 years and did not differ between groups (p=0.77).

**Table 1.** Baseline cohort characteristics.

|  | Active RRMS<br>(n=9) | Stable RRMS<br>(n=33) | PMS<br>(n=17) | P value<br>stable<br>RRMS vs<br>PMS |
| --- | --- | --- | --- | --- |
| Female<sup>1</sup> | 6, 67% | 22, 67% | 11, 65% | >0.99<sup>4</sup> |
| Age (years)<sup>2</sup> | 46 ± 10 | 45 ± 11 | 57 ± 9 | 0.0005<sup>5,#</sup> |
| Disease duration<sup>2</sup> | 11 ± 9 | 11 ± 8 | 23 ± 11 | <0.0001<sup>5,#</sup> |
| 7T follow-up time (years)<sup>2</sup> | 1.9 ± 0.7 | 2.0 ± 0.4 | 1.9 ± 0.6 | 0.40<sup>5</sup> |
| Clinical follow-up time (years)<sup>2</sup> | 2.4 ± 0.9 | 3.0 ± 0.6 | 2.6 ± 0.7 | 0.07<sup>5</sup> |
| Disability measures |  |  |  |  |
| EDSS<sup>3</sup> | 1, 1.5 (0-6.5) | 1.5, 1.0 (0-5.0) | 6, 3 (2-7.5) | <0.0001<sup>6,#</sup> |
| 25FTW<sup>2</sup> | 6.4 ± 6.0 | 4.7 ± 1.0 | 13.9 ± 11.9 | <0.0001<sup>5,#</sup> |
| 9HPT<sup>2</sup> | 20.0 ± 6.3 | 19.3 ± 3.1 | 37.8 ± 17.9 | <0.0001<sup>5,#</sup> |
| PASAT<sup>2</sup> | 49 ± 6 | 53 ± 7 | 43 ± 12 | 0.0004<sup>5,#</sup> |
| SDMT<sup>2</sup> | 61 ± 16 | 58 ± 10 | 35 ± 14 | <0.0001<sup>5,#</sup> |
| Treatment<sup>1</sup> |  |  |  |  |
| None | 5, 56% | 4, 12% | 2, 12% | 0.62<sup>7</sup> |
| Injectable | 2, 22% | 6, 18% | 3, 18% |  |
| Oral | 1, 11% | 15, 34% | 5, 29% |  |
| Infusion | 1, 11% | 8, 24% | 7, 41% |  |
| MRI measures |  |  |  |  |
| Cortical lesions<sup>3</sup> |  |  |  |  |
| Total cortical lesion number | 18, 71 (0-168) | 15, 22 (0-113) | 58, 100 (2-212) | 0.006<sup>6,#</sup> |
| Total cortical lesion volume (μl) | 530, 1488<br>(0-9888) | 375, 903<br>(0-6392) | 1268, 3310<br>(32-8918) | 0.04<sup>6</sup> |
| Leukocortical lesion number | 3, 10 (0-17) | 5, 12 (0-67) | 29, 37 (1-49) | 0.008<sup>6,#</sup> |
| Leukocortical lesion volume (μl) | 48, 165 (0-214) | 122, 368 (0-2818) | 553, 950 (11-1709) | 0.02<sup>6,#</sup> |
| Intracortical lesion number | 1, 7 (0-20) | 1, 2 (0-7) | 4, 8 (0-10) | 0.004<sup>6,#</sup> |
| Intracortical lesion volume (μl) | 3, 26 (0-65) | 4, 8 (0-34) | 11, 30 (0-71) | 0.02<sup>6,#</sup> |
| Subpial lesion number | 17, 49 (0-142) | 5, 10 (0-93) | 21, 52 (0-168) | 0.007<sup>6,#</sup> |
| Subpial lesion volume (μl) | 287, 1426<br>(0-9660) | 143, 429<br>(0-5233) | 505, 2403<br>(0-8015) | 0.02<sup>6,#</sup> |
| White matter lesion volume (ml)<sup>3</sup> | 4.2, 8.0<br>(1.8-26.5) | 6.5, 7.9<br>(0.9-49.9) | 13, 21<br>(1.6-63.1) | 0.01<sup>6,#</sup> |
| Spinal cord lesion number<sup>3</sup> | 4, 6 (0-10) | 5, 7 (0-13) | 9.5, 7 (1-19) | 0.004<sup>6,#</sup> |
| Normalized brain volume (ml)<sup>2</sup> |  |  |  |  |
| Supratentorial brain | 1,261 ± 31 | 1,256 ± 14 | 1,175 ± 23 | 0.003<sup>5,#</sup> |
| Deep gray matter | 50 ± 2 | 48 ± 7 | 45 ± 2 | 0.07<sup>5</sup> |
| Cortex | 614 ± 13 | 611 ± 7 | 589 ± 8 | 0.048<sup>5</sup> |
| White matter | 580 ± 18 | 580 ± 9 | 526 ± 16 | 0.003<sup>5,#</sup> |

RRMS: relapsing remitting multiple sclerosis. Active RRMS was defined as the presence of a new or enhancing lesion on MRI in the year prior to enrollment in the study. PMS: progressive multiple sclerosis. EDSS: expanded disability status scale. 25FTW: 25 foot timed walk. 9HPT: 9-hole peg test. PASAT: paced auditory serial addition test. SDMT: symbol digit modality test. Raw scores are used for all disability measures. Injectable: glatiramer acetate, interferons. Oral: dimethyl fumarate, fingolimod, teriflunomide. Infusion: daclizumab, natalizumab, ocrelizumab, rituximab. <sup>1</sup>n (%). <sup>2</sup>mean ± standard deviation. <sup>3</sup>median, interquartile range (range). <sup>4</sup>Fisher's exact test. <sup>5</sup>t-test. <sup>6</sup>Mann-Whitney test. <sup>7</sup>Chi-square test. #Significant after adjusting for false-discovery rate.

12 individuals formed at least one new cortical lesion, including 5/9 (56%) active RRMS, 3/33 (9%) stable RRMS, and 4/17 (24%) progressive MS (p=0.008, stable RR vs PMS p=0.21). A total of 16 new cortical lesions formed in the active RRMS group (median 1 per participant, IQR 2, range 0-10), 7 in the stable RRMS group (median 0, IQR 0, range 0-5), and 4 in the progressive MS group (median 0, IQR 1, range 0-1) (p=0.006, stable RR vs PMS p=0.88) (Figure 1A, Table 2). There was no difference in the subtypes of new cortical lesions (leukocortical, intracortical, and subpial) between the three groups (p=0.87).

**Figure 1.**
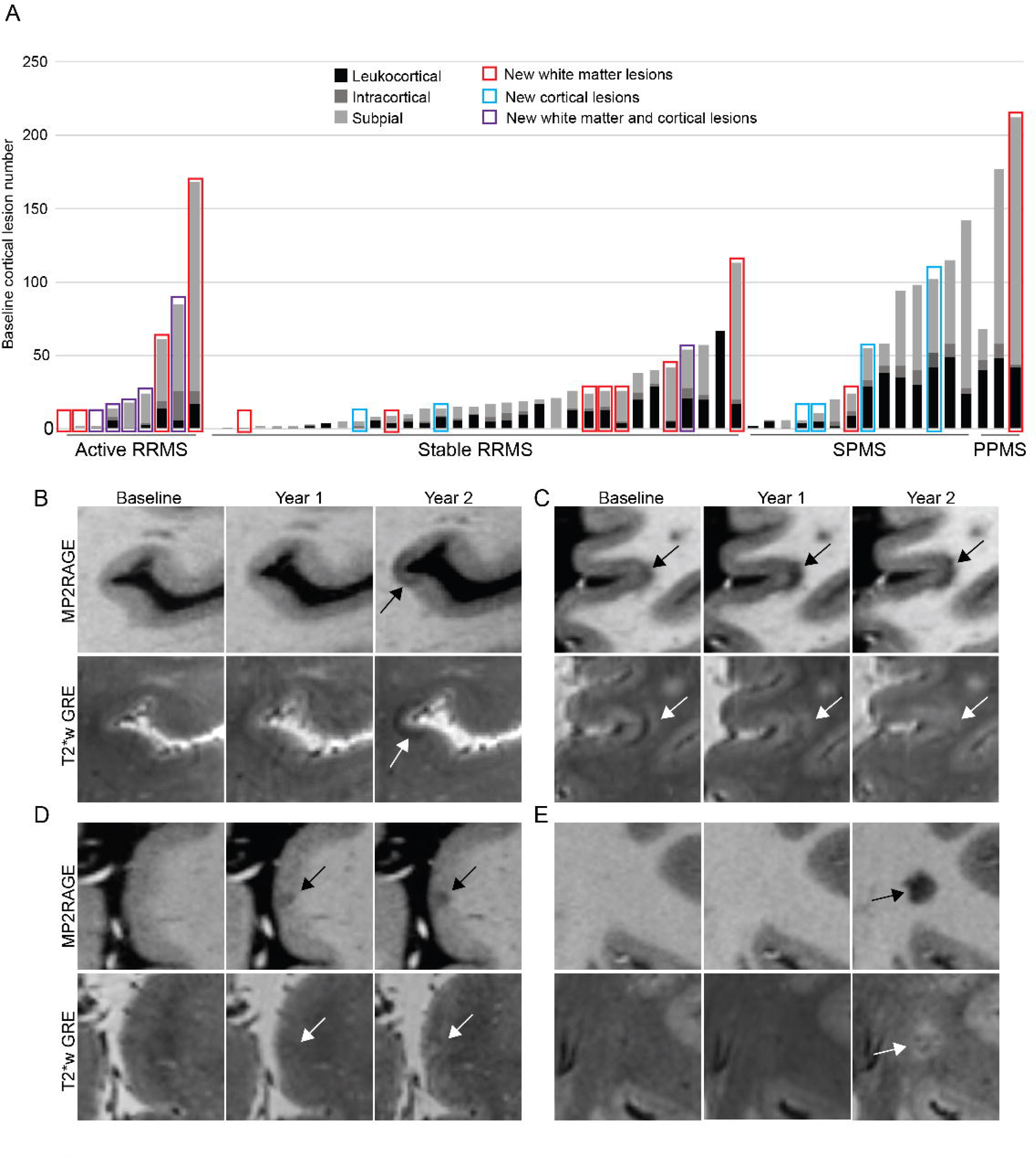
New cortical lesion formation is rare in people with stable white matter lesions. (A) New cortical and white matter lesion formation was frequent in people with active RRMS (defined as having at least one new or enhancing white matter lesion in the year prior to enrollment) but was infrequent in people with radiographically stable RRMS or progressive MS. New cortical lesion formation was not related to baseline cortical lesion number (gray bars, each bar represents a single participant). (B-D) Appearance of new cortical lesions can differ from the appearance of chronic cortical lesions (cortical lesions are denoted with arrows). (B) A subpial lesion that was new on the year-2 MRI appears hypointense on T_2_*w GRE. (C) A leukocortical lesion that was present at baseline is initially hyperintense on T_2_*w GRE with a band of hypointensity at the cortex-white matter junction. This lesion expands over time and becomes more hypointense on T_1_w MP2RAGE, while at the same time the hypointense band on T_2_*w GRE disappears. (D) A subpial lesion that was new on the year-1 MRI initially appears isointense on T_2_*w GRE and then becomes more hyperintense at year-2. (E) A white matter lesion in the same individual as (C) was new on the year-2 MRI and is hyperintense on T_2_*w GRE. RRMS: relapsing remitting multiple sclerosis. SPMS: secondary progressive multiple sclerosis. PPMS: primary progressive multiple sclerosis. T_2_*w GRE: T_2_* weighted gradient recalled echo. MP2RAGE: magnetization prepared 2 rapid acquisition gradient echo.

**Table 2.** Longitudinal MRI changes by MS phenotype.

|  | Active RRMS<br>(n=9) | Stable RRMS<br>(n=33) | PMS (n=17) | P value (Active RRMS<br>vs stable<br>RRMS vs<br>PMS) | P value<br>(Stable<br>RRMS<br>vs PMS) |
| --- | --- | --- | --- | --- | --- |
| **Cortical lesions** |  |  |  |  |  |
| Number of individuals with new lesions<sup>1</sup> | 5 (56%) | 3 (9%) | 4 (24%) | 0.008<sup>5,#</sup> | 0.21<sup>6</sup> |
| New lesion number<sup>2</sup> | 16, 1, 2 (0 – 10) | 7, 0, 0 (0 – 5) | 4, 0, 1 (0 – 1) | 0.006<sup>7,#</sup> | 0.88<sup>8</sup> |
| New lesion volume (μl)<sup>3</sup> | 3.5, 67.75<br>(0 – 484.4) | 0, 0<br>(0 – 51) | 0, 1<br>(0 – 19.25) | 0.020<sup>7</sup> | >0.99<sup>8</sup> |
| Change in lesion volume (μl)<sup>3</sup> | 68.3, 151.7<br>(2.8 – 490.2) | 8.1, 32.6<br>(-60.6 – 154.2) | 22.1, 84.3<br>(-23.5 – 215.3) | 0.046<sup>7</sup> | 0.49<sup>8</sup> |
| % change in lesion volume<sup>3</sup> | 7, 13<br>(1 – 297) | 2, 8<br>(-5 – 155) | 1.4, 4.49<br>(-6.5 – 18.6) | 0.05<sup>7</sup> | >0.99<sup>8</sup> |
| **White matter lesions** |  |  |  |  |  |
| Number of individuals with new lesions<sup>1</sup> | 9 (100%) | 8 (24%) | 2 (17%) | <0.0001<sup>7,#</sup> | 0.46<sup>8</sup> |
| New lesion number<sup>2</sup> | 66, 2, 12 (1 – 31) | 27, 0, 1 (0 – 9) | 3, 0, 0 (0 – 2) | <0.0001<sup>7,#</sup> | >0.99<sup>8</sup> |
| New lesion volume (μl)<sup>3</sup> | 87, 358 (0 – 675) | 0, 8 (0 – 498) | 0, 0 (0 – 14) | <0.0001<sup>7,#</sup> | 0.38<sup>8</sup> |
| Change in lesion volume (μl)<sup>3</sup> | 127, 453<br>(-306 – 2057) | 38, 151.5<br>(-2238 – 903) | 44, 79<br>(-2458 – 1816) | 0.17<sup>7</sup> | >0.99<sup>8</sup> |
| % change in lesion volume<sup>3</sup> | 4, 10<br>(-17 – 14) | 1, 1<br>(-14 – 11) | 0.4, 1<br>(-8.9 – 2.9) | 0.04<sup>7</sup> | >0.99<sup>8</sup> |
| **Spinal cord lesions** |  |  |  |  |  |
| Number of individuals with new lesions<sup>1</sup> | 2 (22%) | 2 (6%) | 0 (0%) | 0.10<sup>7</sup> | 0.54<sup>8</sup> |
| New lesion number<sup>2</sup> | 4, 0, 1 (0 – 2) | 6, 0, 0 (0 – 5) | 0, 0, 0 (0 – 0) | 0.10<sup>7</sup> | >0.99<sup>8</sup> |
| **Annualized atrophy rate (%)** |  |  |  |  |  |
| Supratentorial brain | 0.48 ± 0.27 | 0.17 ± 1.74 | -0.07 ± 0.24 | 0.374<sup>9</sup> | 0.417<sup>10</sup> |
| Deep gray matter | 1.41 ± 0.90 | 0.05 ± 0.25 | 0.13 ± 0.38 | 0.105<sup>9</sup> | 0.867<sup>10</sup> |
| Cortex | 0.09 ± 0.60 | 0.21 ± 0.24 | -0.21 ± 0.36 | 0.656<sup>9</sup> | 0.339<sup>10</sup> |
| White matter | 0.78 ± 0.42 | 0.14 ± 0.20 | 0.06 ± 0.32 | 0.278<sup>9</sup> | 0.829<sup>10</sup> |

RRMS: relapsing remitting multiple sclerosis. Active RRMS was defined as the presence of a new or enhancing lesion on MRI in the year prior to enrollment in the study. PMS: progressive multiple sclerosis. <sup>1</sup>n (%). <sup>2</sup>total, median, interquartile range (range). <sup>3</sup>median, interquartile range (range). <sup>4</sup>mean ± standard deviation. <sup>5</sup>Chi-square. <sup>6</sup>Fisher's exact test. <sup>7</sup>Kruskal-Wallis test. <sup>8</sup>Mann-Whitney test. <sup>9</sup>One-way ANOVA. <sup>10</sup>t-test. #Significant after adjusting for false-discovery rate.

Individuals with new cortical lesions did not differ from those with no new cortical lesions with respect to age, sex, disease duration, or baseline cortical lesion burden (Table 3). Disease modifying therapy (categorized as none, injectable, oral, infusion, or changed during the study) did not differ between people with new vs no new cortical lesions. 3/9 people on infusion therapies formed one new cortical lesion each. Two people were on ocrelizumab: one received their first dose approximately 1 year before the baseline MRI and the other received their first dose 8 months before the baseline MRI; neither had any interruptions in treatment. One person was on natalizumab for over 1 year before the baseline MRI and did not have any known interruptions in treatment. There was no difference in change in any of the disability measures or in overall disability progression (defined as an increase in EDSS of ≥1 for baseline EDSS <6 or ≥0.5 for baseline EDSS ≥6 or 20% increase in 25TW or 20% increase in 9HPT) in people with new vs no new cortical lesions (mean clinical follow-up time 2.8 ± 0.7 years).

**Table 3.** Characteristics of individuals with new vs stable lesions.

|  | Cortical lesions |  |  | White matter lesions |  |  |
| --- | --- | --- | --- | --- | --- | --- |
|  | New lesions (n=12) | No new lesions (n=47) | P value | New lesions (n=19) | No new lesions (n=40) | P value |
| Age (years)<sup>1</sup> | 48 ± 11 | 49 ± 11 | 0.92<sup>7</sup> | 47 ± 10 | 50 ± 12 | 0.32<sup>7</sup> |
| Female<sup>2</sup> | 10 (83%) | 29 (62%) | 0.19<sup>8</sup> | 14 (74%) | 25 (63%) | 0.56<sup>8</sup> |
| Disease duration (years)<sup>1</sup> | 13 ± 11 | 15 ± 11 | 0.71<sup>7</sup> | 12 ± 10 | 15 ± 11 | 0.22<sup>7</sup> |
| Baseline lesions |  |  |  |  |  |  |
| Cortical lesion number<sup>3</sup> | 16, 48<br>(2 – 102) | 20, 53<br>(0-212) | 0.97<sup>9</sup> | 24, 52<br>(0 – 212) | 16, 52<br>(0 – 177) | 0.37<sup>9</sup> |
| Cortical lesion volume (μl)<sup>3</sup> | 445, 1072<br>(23 – 3702) | 384, 2059<br>(0 – 9888) | 0.99<sup>9</sup> | 782, 1960<br>(0-9888) | 367, 1231<br>(0 – 5159) | 0.43<sup>9</sup> |
| White matter lesion volume (ml)<sup>3</sup> | 7.2, 11.7<br>(1.8 – 32.9) | 8.4, 10.2<br>(0.9 – 63.1) | 0.92<sup>9</sup> | 5.3, 8.5<br>(1.8 – 26.5) | 8.5, 14.2<br>(0.9 – 62.2) | 0.22<sup>9</sup> |
| Treatment<sup>4</sup> |  |  |  |  |  |  |
| None | 2 (17%) | 4 (9%) | 0.10<sup>10</sup> | 3 (16%) | 3 (8%) | 0.42<sup>10</sup> |
| Injectable | 3 (25%) | 5 (11%) |  | 2 (11%) | 6 (15%) |  |
| Oral | 0 (0%) | 14 (30%) |  | 4 (21%) | 10 (25%) |  |
| Infusion | 3 (25%) | 6 (13%) |  | 1 (5%) | 8 (20%) |  |
| Switch | 4 (33%) | 18 (38%) |  | 9 (47%) | 13 (33%) |  |
| Change in disability |  |  |  |  |  |  |
| Change in EDSS<sup>3</sup> | 0, 0.5<br>(-1 – 0.5) | 0, 1<br>(-1 – 4) | 0.43<sup>9</sup> | 0, 1<br>(-1 – 2) | 0, 1<br>(-1 – 4) | 0.27<sup>9</sup> |
| 25FTW % change<sup>1</sup> | 18 ± 38 | 23 ± 51 | 0.75<sup>7</sup> | 8 ± 18 | 28 ± 56 | 0.11<sup>7</sup> |
| 9HPT % change<sup>1,5</sup> | 18 ± 37 | 15 ± 32 | 0.78<sup>7</sup> | 10 ± 23 | 18 ± 36 | 0.40<sup>7</sup> |
| PASAT % change<sup>1</sup> | 5 ± 10 | -1 ± 14 | 0.19<sup>7</sup> | 2 ± 10 | -1 ± 13 | 0.49<sup>7</sup> |
| SDMT % change<sup>1</sup> | -3 ± 13 | 1 ± 17 | 0.50<sup>7</sup> | 3 ± 20 | -2 ± 13 | 0.37<sup>7</sup> |
| Overall disability progression<sup>6</sup> | 3 (25%) | 23 (49%) | 0.20<sup>8</sup> | 5 (26%) | 22 (55%) | 0.05<sup>8</sup> |
| Transition from RRMS to SPMS<sup>2</sup> | 0 (0%) | 5 (15%) | 0.56<sup>8</sup> | 3 (18%) | 2 (8%) | 0.38<sup>8</sup> |

EDSS: expanded disability status scale. 25FTW: 25 foot timed walk. 9HPT: 9-hole peg test. PASAT: paced auditory serial addition test. SDMT: symbol digit modality test. RRMS: relapsing remitting multiple sclerosis. PMS: progressive multiple sclerosis. <sup>1</sup>mean ± standard deviation. <sup>2</sup>n (%). <sup>3</sup>median, interquartile range (range). <sup>4</sup>Injectable: glatiramer acetate, interferons. Oral: dimethyl fumarate, fingolimod, teriflunomide. Infusion: daclizumab, natalizumab, ocrelizumab, rituximab. <sup>5</sup>In hand with greater worsening. <sup>6</sup>Overall disability progression defined as an increase in EDSS of ≥1 for baseline EDSS <6 or ≥0.5 for baseline EDSS ≥6 or 20% increase in 25TW or 20% increase in 9HPT. <sup>7</sup>t-test. <sup>8</sup>Fisher's exact test. <sup>9</sup>Mann-Whitney test. <sup>10</sup>Chi-square test.

19 individuals formed a total of 96 new white matter lesion, including 9/9 (100%) active RRMS, 8/33 (24%) stable RRMS, and 2/17 (12%) progressive MS (p<0.0001, stable RR vs PMS p=0.46) (Figure 1A, Table 2). There were no differences in age, sex, disease duration, baseline white matter lesion volume, disease modifying therapy, or change in disability between people with new vs no new white matter lesions. 1/9 people on infusion therapies (ocrelizumab) formed a new white matter lesion; this individual’s first dose of ocrelizumab was two months before their baseline MRI. There was no difference in change in any of the disability measures or in overall disability progression in people with new vs no new white matter lesions (Table 3).

6/12 people (50%) with new cortical lesions also formed new white matter lesions, while 13/47 (27%) of people without new cortical lesions formed new white matter lesions (p=0.17). There was no difference in the subtypes of cortical lesions that formed in people with vs without new white matter lesions (p=0.69). Three white matter lesions in three individuals expanded into the cortex during the 7T follow up period. No intracortical or subpial lesions expanded into the white matter.

New spinal cord lesions were rare (10 total new lesions in 4 individuals) and did not differ between groups, nor did change in brain volume (Table 2).

Four individuals had a total of six relapses during the follow-up period. Two individuals were in the active RRMS group, and both had new lesions in the white matter, spinal cord, and cortex. Two individuals with clinical relapses were in the stable RRMS group; neither of these individuals had new lesions in the brain or spinal cord. None of the four had overall disability progression (composite measure of EDSS, 9HPT, and 25TW), and none transitioned from RRMS to SPMS.

### New cortical lesions have a distinct appearance on T_2_*w images

While most cortical lesions are hypointense on T_1_w images and hyperintense on T_2_*w images, we observed that 35% of the new cortical lesions, while hypointense on T_1_w images, were hypointense or isointense on T_2_*w images. In contrast, only 10% of new white matter lesions were iso- or hypointense on T_2_*w images (p=0.01) (Figure 1B-E).

### Baseline cortical lesion burden, but not white matter or spinal cord lesion burden, is associated with worsening disability and transition from relapsing to progressive MS

To determine the association between baseline cortical lesion burden and subsequent change in disability, we considered all available timepoints at which there was clinical follow-up. Mean ± SD clinical follow up time was 2.8 ± 0.7 years. There was no difference in clinical follow-up time between active RRMS, stable RRMS, and progressive MS (p=0.07).

Baseline total cortical lesion volume was correlated with percent change in 9HPT (r=0.69, p<0.001, adjusted for age and sex, significant after adjusting for multiple comparisons) and 25FTW (r=0.49, p<0.001, significant after adjusting for multiple comparisons). Baseline leukocortical, intracortical, and subpial lesion volume were each associated with percent change in 9HPT and percent change in 25FTW. Baseline cortical lesion volume was not correlated with change in EDSS, PASAT, or SDMT. Baseline white matter lesion volume was correlated with percent change in 25FTW (r=0.45, p<0.001) and PASAT (r=-0.38, p<0.01). Baseline spinal cord lesion number and normalized brain volume were not correlated with change in any of the individual disability measures (Table 4).

**Table 4.**
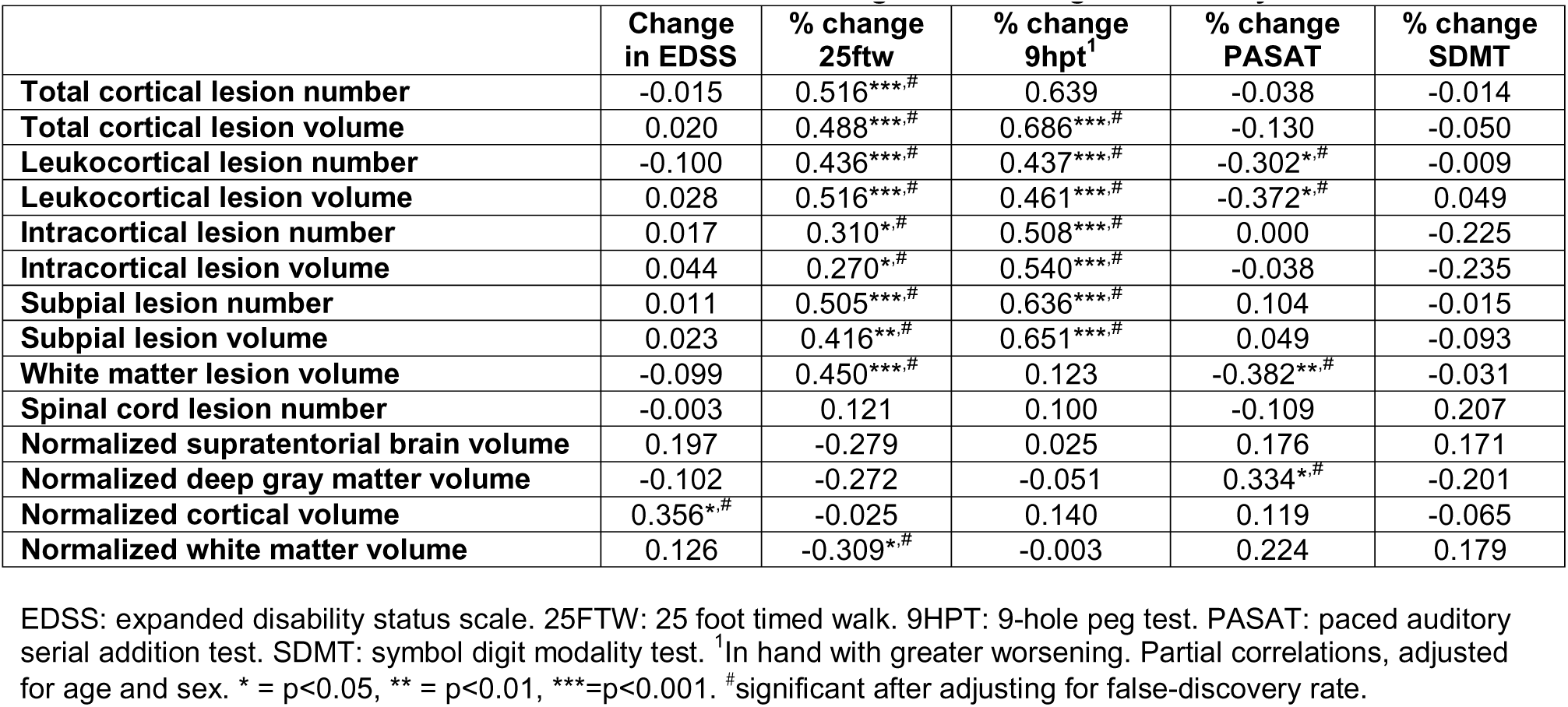
Association between baseline MRI measures and longitudinal change in disability.

We also determined whether baseline lesion burden was associated with overall progression of disability, defined as an increase in EDSS of ≥1 for baseline EDSS <6 or ≥0.5 for baseline EDSS ≥6 or 20% increase in 25TW or 20% increase in 9HPT. Baseline cortical lesion volume was higher in people with subsequent progression of disability (median 1010 ml, IQR 3162, range 13-9888) than in those without (median 267 ml, IQR 854, range 0-3539, B 0.809, p=0.001, adjusted for age and sex) (Figure 2A, table 5). Baseline volume was also higher in people with progression disability for each of the cortical lesion subtypes. In contrast, baseline white matter lesion volume did not differ between people with subsequent progression of disability (median 10.4 ml, IQR 14.2, range 1.5-63.1) vs those without (median 6.5 ml, IQR 12.0, range 0.9-50.8, B 0.206, p=0.42). There was also no difference in baseline spinal cord lesion number or baseline normalized brain volume between people with progression of disability vs those without (Figure 2A, table 5).

**Figure 2.**
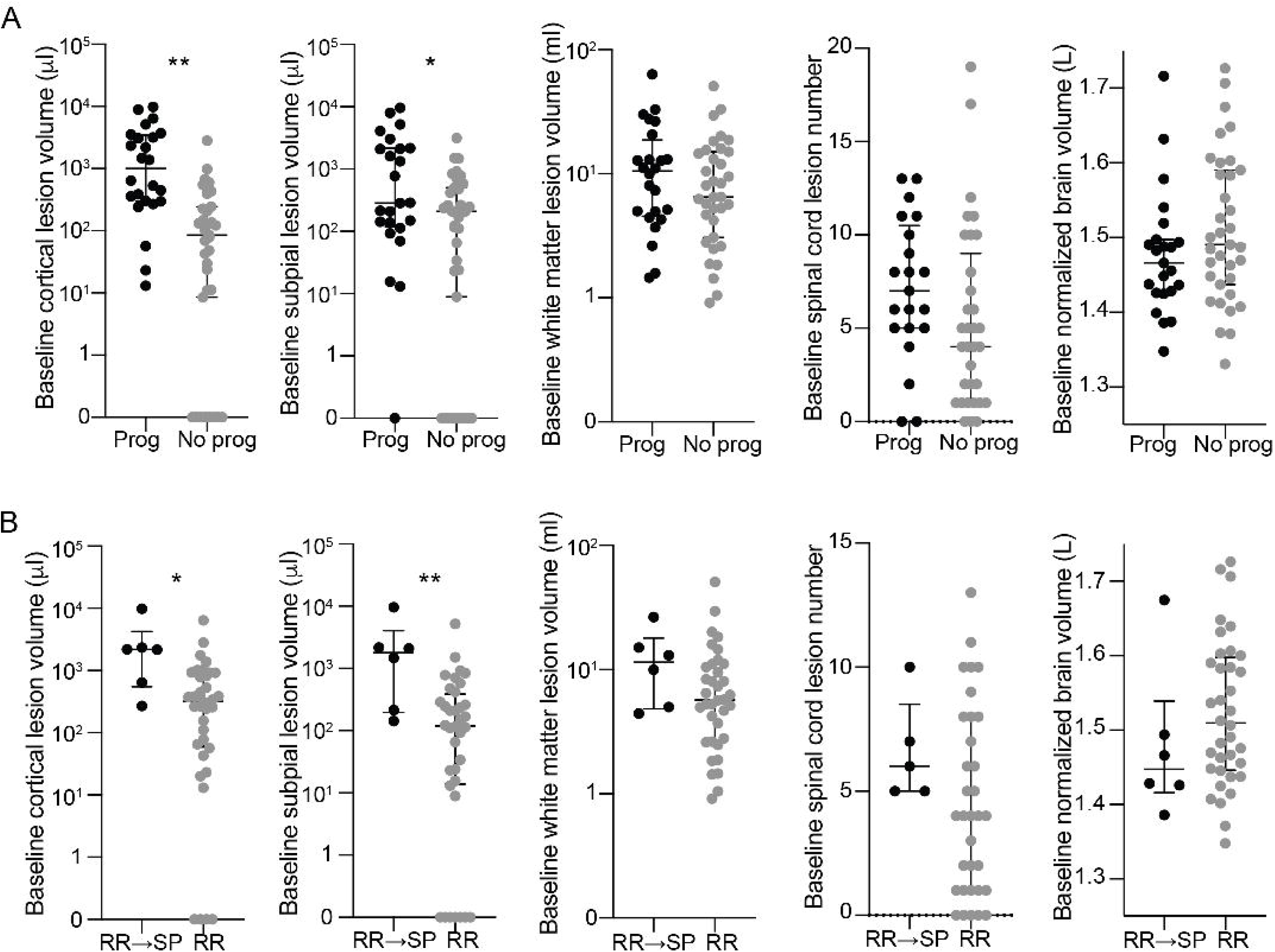
People with worsening disability and transition from relapsing to progressive MS have higher cortical lesion burden at baseline. (A) Individuals with progression of disability during the study (“Prog,” defined as an increase in EDSS of ≥1 for baseline EDSS <6 or ≥0.5 for baseline EDSS ≥6 or 20% increase in 25TW or 20% increase in 9HPT) had higher baseline cortical lesion volume and subpial lesion volume, but no difference in baseline white matter lesion volume, spinal cord lesion number, or normalized brain volume, compared to people without progression of disability (“No prog”). Similar results were observed when comparing individuals who transitioned from relapsing remitting to secondary progressive MS with those who remained relapsing remitting (B). Comparisons were performed using a multivariable generalized linear model adjusted for age and sex. RR: relapsing remitting. SP: secondary progressive. * p<0.05, ** p<0.01

**Table 5.**
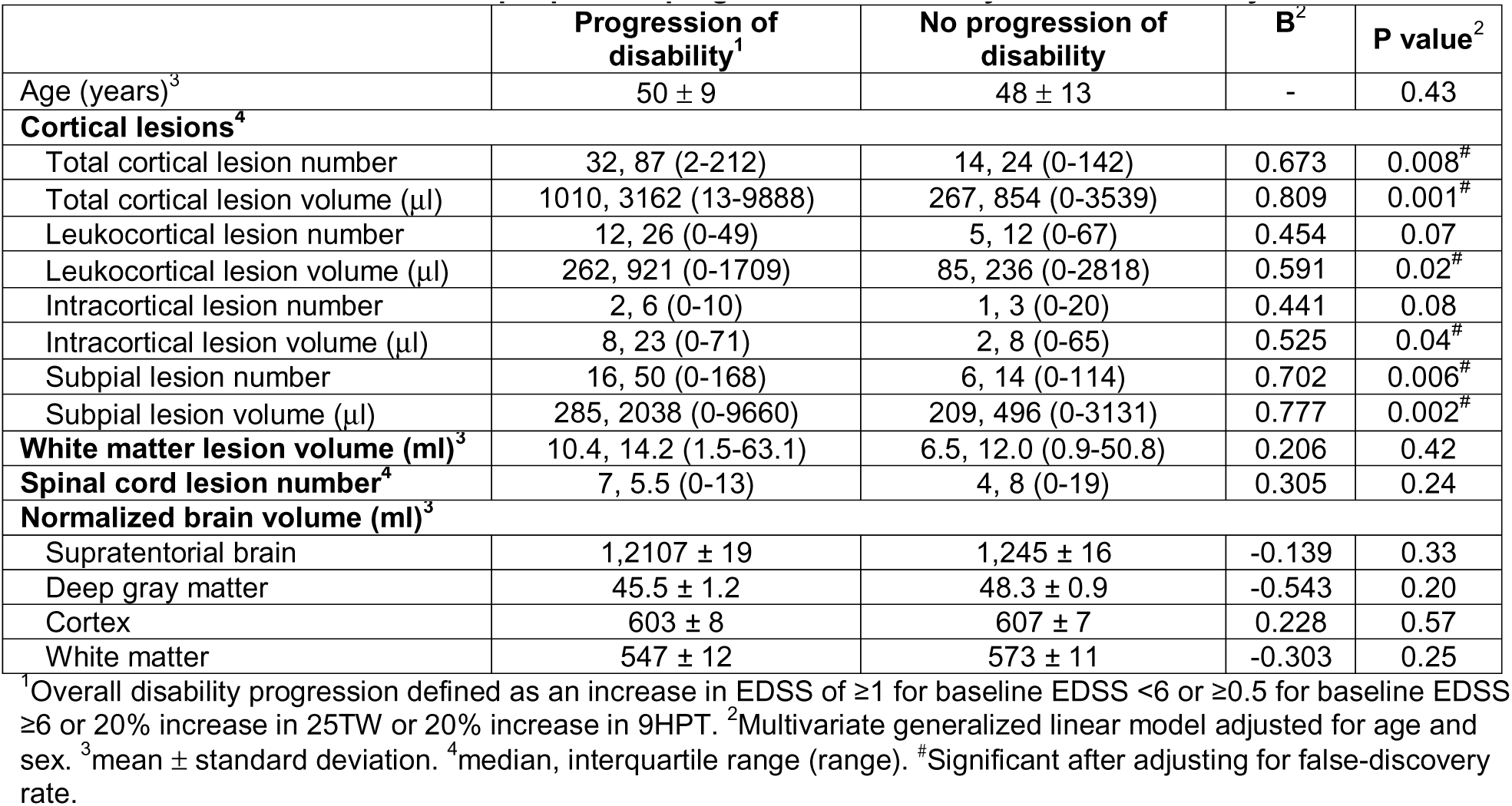
Baseline MRI measures in people with progression of disability vs stable disability.

|  | **Progression of disability<sup>1</sup>** | **No progression of disability** | **B<sup>2</sup>** | **P value<sup>2</sup>** |
| --- | --- | --- | --- | --- |
| Age (years)<sup>3</sup> | 50 ± 9 | 48 ± 13 | - | 0.43 |
| **Cortical lesions<sup>4</sup>** |  |  |  |  |
| Total cortical lesion number | 32, 87 (2-212) | 14, 24 (0-142) | 0.673 | 0.008<sup>#</sup> |
| Total cortical lesion volume (μl) | 1010, 3162 (13-9888) | 267, 854 (0-3539) | 0.809 | 0.001<sup>#</sup> |
| Leukocortical lesion number | 12, 26 (0-49) | 5, 12 (0-67) | 0.454 | 0.07 |
| Leukocortical lesion volume (μl) | 262, 921 (0-1709) | 85, 236 (0-2818) | 0.591 | 0.02<sup>#</sup> |
| Intracortical lesion number | 2, 6 (0-10) | 1, 3 (0-20) | 0.441 | 0.08 |
| Intracortical lesion volume (μl) | 8, 23 (0-71) | 2, 8 (0-65) | 0.525 | 0.04<sup>#</sup> |
| Subpial lesion number | 16, 50 (0-168) | 6, 14 (0-114) | 0.702 | 0.006<sup>#</sup> |
| Subpial lesion volume (μl) | 285, 2038 (0-9660) | 209, 496 (0-3131) | 0.777 | 0.002<sup>#</sup> |
| **White matter lesion volume (ml)<sup>3</sup>** | 10.4, 14.2 (1.5-63.1) | 6.5, 12.0 (0.9-50.8) | 0.206 | 0.42 |
| **Spinal cord lesion number<sup>4</sup>** | 7, 5.5 (0-13) | 4, 8 (0-19) | 0.305 | 0.24 |
| **Normalized brain volume (ml)<sup>3</sup>** |  |  |  |  |
| Supratentorial brain | 1,2107 ± 19 | 1,245 ± 16 | -0.139 | 0.33 |
| Deep gray matter | 45.5 ± 1.2 | 48.3 ± 0.9 | -0.543 | 0.20 |
| Cortex | 603 ± 8 | 607 ± 7 | 0.228 | 0.57 |
| White matter | 547 ± 12 | 573 ± 11 | -0.303 | 0.25 |

<sup>1</sup>Overall disability progression defined as an increase in EDSS of ≥1 for baseline EDSS <6 or ≥0.5 for baseline EDSS ≥6 or 20% increase in 25TW or 20% increase in 9HPT. <sup>2</sup>Multivariate generalized linear model adjusted for age and sex. <sup>3</sup>mean ± standard deviation. <sup>4</sup>median, interquartile range (range). <sup>#</sup>Significant after adjusting for false-discovery rate.

Increase in cortical lesion volume was higher in people with progression of disability (B 0.831, p=0.007, adjusted for age and sex), however this was not the case for percent change in cortical lesion volume, nor was change in cortical lesion volume associated with progression of disability when baseline cortical lesion volume was included in the model (B -0.245, p=0.56). Change in white matter lesion volume and rate of brain atrophy did not differ between people with progression of disability vs those without (Supplementary table 2).

6/42 individuals with RRMS at baseline had transitioned to SPMS by the time of the final clinical follow-up, as determined clinically by the evaluating neurologist. At baseline, individuals who subsequently transitioned to SPMS had higher total cortical lesion volume (median 2183 μl, IQR 3673, range 270-9888 vs median 321 μl, IQR 832, range 0-6392, B 0.873, p=0.01, adjusted for age and sex), and subpial lesion volume (median 1489 μl, IQR 3850, range 143-9660 vs median 119 μl, IQR 372, range 0-5233, B=1.047, p=0.003) than those who remained RRMS. There was no difference in leukocortical lesion volume, white matter lesion volume, or spinal cord lesion number between those who transitioned from RRMS to SPMS vs those who remained relapsing remitting, nor did baseline brain volume differ between the groups (Figure 2B, Supplementary Table 3).

### Association between baseline lesion burden and rate of subsequent brain atrophy

Baseline white matter lesion volume was associated with subsequent brain atrophy rate (r=0.320, p=0.04, adjusted for age and sex), as was leukocortical lesion number (r=0.375, p=0.02). There was no association between baseline cortical lesion number or volume, subpial or intracortical lesion number or volume, or spinal cord lesion number and subsequent change in brain volumes (Supplementary Table 4).

## Discussion

We characterized cortical lesions longitudinally in a cohort of individuals with MS, most of whom had not formed new white matter lesions in the year prior to enrollment in the study. We found that cortical lesion formation was rare in this population, was not higher in progressive vs relapsing MS, and was not associated with worsening of disability over time. Importantly, however, baseline cortical lesion burden, but not white matter or spinal cord lesion burden, was higher in people who had subsequent worsening of motor disability and in people who transitioned from relapsing to secondary progressive MS.

Cortical lesion formation in this study was lower than what has previously been reported,^29, 30^ especially in the participants who had stable white matter lesions prior to enrollment. Also in contrast to prior studies,^13, 14^ we did not find a higher rate of cortical lesion formation in progressive MS. These previous studies were published between 2009 and 2019, and participants were on lower efficacy treatments than the participants here, of whom 37/59 (63%) were on oral or infusion disease modifying therapies. Limited observation has previously demonstrated lower rates of cortical lesion formation in people on disease-modifying therapy.^31, 32^ However, these studies were done using 1.5T double inversion recovery MRI, which has poor sensitivity for cortical, and in particular subpial, lesions.^33^ Dedicated prospective studies will be needed to test the effects of modern disease-modifying therapies on cortical lesion formation. It is possible that despite evident differences in mechanisms of cortical and white matter lesion formation within the central nervous system, peripheral immune mediators targeted by disease-modifying therapies may be shared. The lower rate of cortical lesion formation in this cohort could also be due to the purposeful recruitment of individuals with stable white matter lesions, resulting in a cohort that was older (mean 49 years) than in prior studies and for the most part had longstanding disease (mean 14 years). We did not find differences in baseline demographic, clinical, or MRI measures in individuals with new vs no new cortical lesions, however this may be due to the very small number of individuals with new cortical lesions.

Our finding that cortical lesion burden predicts disability progression and transition from relapsing to secondary progressive MS is in agreement with prior studies, including at least one prior 7T study.^14, 30, 34, 35^ The lack of new cortical lesions in this older cohort with longstanding disease, combined with the association between baseline cortical lesion burden and 2-year disability worsening, suggests that, at least in the current era, cortical lesions form early in disease^12, 36^ and then may exert long-term effects on disability.

Cortical lesions could potentially exert these long term effects on disability by causing local and/or remote neurodegeneration and cell death, reflected as decreased brain volume on MRI. In both cross-sectional and longitudinal studies, the relationship between cortical lesion burden and whole brain and cortical atrophy has been inconsistent.^6, 23, 34, 37–39^ Here, we did not find a consistent relationship between baseline cortical lesion burden and subsequent brain atrophy. It is possible that there are more local effects on brain structure or microstructural changes that are not visible with the MRI methods used here.

One potential limitation of this study is that there were changes to scanner software versions on both the 3T and 7T scanners during the study as well as a switch partway through year 2 to a motion and B_0_-corrected version of the T2*w GRE sequence. Because of significant differences in image contrast on 3T MP2RAGE images acquired with different WIP sequences, which we found affected tissue segmentation and brain volume calculations, we used 7T MP2RAGE images for brain volume calculations. This meant that we were unable to include the infratentorial brain in our atrophy measurements due to inconsistent coverage. In addition, the lack of a longitudinal brain segmentation methods that works well with 7T images meant that we instead used a cross-sectional method at each timepoint, potentially decreasing our power to detect changes in brain volume over time. For longitudinal cortical lesion assessment, we were careful not to count as “new” any cortical lesion that was clearly seen on the motion corrected T_2_*w GRE images but could have been obscured by artifact on images from the earlier timepoints. This occurred rarely, and we did not observe a difference in new cortical lesion count between timepoints with vs without motion correction, but it is possible that there was a slight undercounting of new cortical lesions.

This study is also limited by cohort size, which, although larger than most 7T studies in MS to date, limits our ability to identify factors associated with cortical lesion formation. This is an important question for future studies, as cortical and white matter lesion burden are not well correlated and the factors determining which patients develop cortical vs white matter lesions are unclear.

If confirmed, our results have important implications for clinical care and research. They suggest that it is necessary to stop cortical lesion formation early in disease and provide some support for the possibility that existing disease-modifying therapies may offer at least partial control. The association we find between cortical lesion burden and subsequent worsening of disability may be useful for prognostication and for selecting participants for clinical trials in progressive MS who are likely to worsen over a relatively short follow up and might benefit most from treatment.

## Data Availability

All data produced in the present study are available upon reasonable request to the authors

## Acknowledgments

We acknowledge the staff of the National Institute of Neurological Disorders and Stroke Neuroimmunology Clinic for care of and collection of clinical data from the study participants and the staff of the National Institute of Mental Health Functional MRI Facility, Matthew Schindler, Martina Absinta, Hadar Kolb, and Maxime Donadieu for help with MRI acquisitions. We thank Siemens for providing access to research pulse sequences. We thank Tianxia Wu for her advice on statistical analyses.

## References

1. Kuhlmann T, Moccia M, Coetzee T, et al. Multiple sclerosis progression: time for a new mechanism-driven framework. Lancet Neurol. Jan 2023;22(1):78–88. doi:10.1016/S1474-4422(22)00289-7

2. Reich DS, Lucchinetti CF, Calabresi PA. Multiple Sclerosis. N Engl J Med. Jan 11 2018;378(2):169–180. doi:10.1056/NEJMra1401483

3. Calabrese M, Agosta F, Rinaldi F, et al. Cortical lesions and atrophy associated with cognitive impairment in relapsing-remitting multiple sclerosis. Arch Neurol. Sep 2009;66(9):1144–50. doi:10.1001/archneurol.2009.174

4. Roosendaal SD, Moraal B, Pouwels PJ, et al. Accumulation of cortical lesions in MS: relation with cognitive impairment. Mult Scler. Jun 2009;15(6):708–14. doi:10.1177/1352458509102907

5. Nielsen AS, Kinkel RP, Madigan N, Tinelli E, Benner T, Mainero C. Contribution of cortical lesion subtypes at 7T MRI to physical and cognitive performance in MS. Neurology. Aug 2013;81(7):641–9. doi:10.1212/WNL.0b013e3182a08ce8

6. Harrison DM, Roy S, Oh J, et al. Association of Cortical Lesion Burden on 7-T Magnetic Resonance Imaging With Cognition and Disability in Multiple Sclerosis. JAMA Neurol. Sep 2015;72(9):1004–12. doi:10.1001/jamaneurol.2015.1241

7. Howell OW, Reeves CA, Nicholas R, et al. Meningeal inflammation is widespread and linked to cortical pathology in multiple sclerosis. Brain. Sep 2011;134(Pt 9):2755–71. doi:10.1093/brain/awr182

8. Magliozzi R, Howell OW, Nicholas R, et al. Inflammatory intrathecal profiles and cortical damage in multiple sclerosis. Ann Neurol. Apr 2018;83(4):739–755. doi:10.1002/ana.25197

9. Magliozzi R, Howell OW, Reeves C, et al. A Gradient of neuronal loss and meningeal inflammation in multiple sclerosis. Ann Neurol. Oct 2010;68(4):477–93. doi:10.1002/ana.22230

10. Bo L, Vedeler CA, Nyland HI, Trapp BD, Mork SJ. Subpial demyelination in the cerebral cortex of multiple sclerosis patients. J Neuropathol Exp Neurol. Jul 2003;62(7):723–32.

11. Brownell B, Hughes JT. The distribution of plaques in the cerebrum in multiple sclerosis. J Neurol Neurosurg Psychiatry. Nov 1962;25:315–20.

12. Lucchinetti CF, Popescu BF, Bunyan RF, et al. Inflammatory cortical demyelination in early multiple sclerosis. N Engl J Med. Dec 08 2011;365(23):2188–97. doi:10.1056/NEJMoa1100648

13. Sethi V, Yousry T, Muhlert N, et al. A longitudinal study of cortical grey matter lesion subtypes in relapse-onset multiple sclerosis. J Neurol Neurosurg Psychiatry. Jul 2016;87(7):750–3. doi:10.1136/jnnp-2015-311102

14. Treaba CA, Granberg TE, Sormani MP, et al. Longitudinal Characterization of Cortical Lesion Development and Evolution in Multiple Sclerosis with 7.0-T MRI. Radiology. 06 2019;291(3):740–749. doi:10.1148/radiol.2019181719

15. Liu J, Beck ES, Filippini S, et al. Navigator-Guided Motion and B0 Correction of T2*-Weighted Magnetic Resonance Imaging Improves Multiple Sclerosis Cortical Lesion Detection. Invest Radiol. Jul 1 2021;56(7):409–416. doi:10.1097/RLI.0000000000000754

16. Liu J, van Gelderen P, de Zwart JA, Duyn JH. Reducing motion sensitivity in 3D high-resolution T2*-weighted MRI by navigator-based motion and nonlinear magnetic field correction. Neuroimage. Feb 1 2020;206:116332. doi:10.1016/j.neuroimage.2019.116332

17. Beck ES, Sati P, Sethi V, et al. Improved Visualization of Cortical Lesions in Multiple Sclerosis Using 7T MP2RAGE. AJNR Am J Neuroradiol. Feb 8 2018;doi:10.3174/ajnr.A5534

18. Jenkinson M, Smith S. A global optimisation method for robust affine registration of brain images. Med Image Anal. Jun 2001;5(2):143–56. doi:10.1016/s1361-8415(01)00036-6

19. Jenkinson M, Bannister P, Brady M, Smith S. Improved optimization for the robust and accurate linear registration and motion correction of brain images. Neuroimage. Oct 2002;17(2):825–41. doi:10.1016/s1053-8119(02)91132-8

20. Cox RW. AFNI: software for analysis and visualization of functional magnetic resonance neuroimages. Comput Biomed Res. Jun 1996;29(3):162–73. doi:10.1006/cbmr.1996.0014

21. Cox RW, Hyde JS. Software tools for analysis and visualization of fMRI data. NMR Biomed. Jun-Aug 1997;10(4-5):171–8. doi:10.1002/(sici)1099-1492(199706/08)10:4/5<171::aid-nbm453>3.0.co;2-l

22. Maranzano J, Dadar M, Rudko DA, et al. Comparison of Multiple Sclerosis Cortical Lesion Types Detected by Multicontrast 3T and 7T MRI. AJNR Am J Neuroradiol. Jul 2019;40(7):1162–1169. doi:10.3174/ajnr.A6099

23. Beck ES, Maranzano J, Luciano NJ, et al. Cortical lesion hotspots and association of subpial lesions with disability in multiple sclerosis. Mult Scler. Aug 2022;28(9):1351–1363. doi:10.1177/13524585211069167

24. Diekhaus H, Donnay C, Gaitan MI, et al. Pseudo-Label Assisted Nnu-Net (PLAn) Enables Automatic Segmentation of 7T MRI From a Single Acquisition. medRxiv. 2022;2022.12.22.22283866 10.1101/2022.12.22.22283866

25. Selvaganesan K, Whitehead E, DeAlwis PM, et al. Robust, atlas-free, automatic segmentation of brain MRI in health and disease. Heliyon. Feb 2019;5(2):e01226. doi:10.1016/j.heliyon.2019.e01226

26. Chernick MR. Bootstrap methods: A guide for practicioners and researchers. 2 ed. Wiley-Interscience; 2008.

27. Efron B. Bootstrap Methods: Another Look at the Jackknife. Annals of Statistics. 1979;7(1):1–26.

28. Benjamini Y, Hochberg Y. Controlling the False Discovery Rate: A Practical and Powerful Approach to Multiple Testing. Journal of the Royal Statistical Society, Series B (Methodological). 1995;57(1):289–300.

29. Calabrese M, Rocca MA, Atzori M, et al. Cortical lesions in primary progressive multiple sclerosis: a 2-year longitudinal MR study. Neurology. Apr 14 2009;72(15):1330–6. doi:10.1212/WNL.0b013e3181a0fee5

30. Calabrese M, Rocca MA, Atzori M, et al. A 3-year magnetic resonance imaging study of cortical lesions in relapse-onset multiple sclerosis. Ann Neurol. Mar 2010;67(3):376–83. doi:10.1002/ana.21906

31. Rinaldi F, Calabrese M, Seppi D, Puthenparampil M, Perini P, Gallo P. Natalizumab strongly suppresses cortical pathology in relapsing-remitting multiple sclerosis. Mult Scler. Dec 2012;18(12):1760–7. doi:10.1177/1352458512447704

32. Rinaldi F, Perini P, Atzori M, Favaretto A, Seppi D, Gallo P. Disease-modifying drugs reduce cortical lesion accumulation and atrophy progression in relapsing-remitting multiple sclerosis: results from a 48-month extension study. Mult Scler Int. 2015;2015:369348. doi:10.1155/2015/369348

33. Kilsdonk ID, Jonkman LE, Klaver R, et al. Increased cortical grey matter lesion detection in multiple sclerosis with 7 T MRI: a post-mortem verification study. Brain. May 2016;139(Pt 5):1472–81. doi:10.1093/brain/aww037

34. Calabrese M, Poretto V, Favaretto A, et al. Cortical lesion load associates with progression of disability in multiple sclerosis. Brain. Oct 2012;135(Pt 10):2952–61. doi:10.1093/brain/aws246

35. Calabrese M, Romualdi C, Poretto V, et al. The changing clinical course of multiple sclerosis: a matter of gray matter. Ann Neurol. Jul 2013;74(1):76–83. doi:10.1002/ana.23882

36. Maranzano J, Till C, Assemlal HE, et al. Detection and clinical correlation of leukocortical lesions in pediatric-onset multiple sclerosis on multi-contrast MRI. Mult Scler. Jun 2019;25(7):980–986. doi:10.1177/1352458518779952

37. Treaba CA, Herranz E, Barletta VT, et al. The relevance of multiple sclerosis cortical lesions on cortical thinning and their clinical impact as assessed by 7.0-T MRI. J Neurol. Jul 2021;268(7):2473–2481. doi:10.1007/s00415-021-10400-4

38. Nielsen AS, Kinkel RP, Tinelli E, Benner T, Cohen-Adad J, Mainero C. Focal cortical lesion detection in multiple sclerosis: 3 Tesla DIR versus 7 Tesla FLASH-T2. J Magn Reson Imaging. Mar 2012;35(3):537–42. doi:10.1002/jmri.22847

39. Pareto D, Sastre-Garriga J, Auger C, et al. Juxtacortical Lesions and Cortical Thinning in Multiple Sclerosis. AJNR Am J Neuroradiol. Dec 2015;36(12):2270–6. doi:10.3174/ajnr.A4485

