## Supplemental tables for "Cortical lesions uniquely predict motor disability accrual and form rarely in the absence of new white matter lesions in multiple sclerosis"

**Supplementary Table 1. MRI sequence parameters.**

|  | **Acquisition plane** | **3D vs 2D** | **TR (ms)** | **TI (ms)** | **TE (ms)** | **FA (°)** | **AT (min:sec)** | **FOV (mm,**  **AP x RL X SI)** | **In-plane resolution (mm)** | **Slice thickness (mm)** |
| --- | --- | --- | --- | --- | --- | --- | --- | --- | --- | --- |
| **3T brain^1^** | | | | | | | | | | |
| PD/T_2_w | Axial | 2D | 5000 | NA | 18, 82 | 150 | 4:25 | 218 x 184 x 138 | 0.34x0.34 | 3 |
| MP2RAGE | Sagittal | 3D | 5000 | 700, 2500 | 2.9 | 4, 5 | 8:16 | 240 x 176 x 256 | 1x1 | 1 |
| FLAIR^2^ | Sagittal | 3D | 4800 | 1800 | 352 | 120 | 6:57 | 256 x 176 x 256 | 1x1 | 1 |
| **3T cervical spine^3^** | | | | | | | | | | |
| STIR | Sagittal | 2D | 8130 | 210 | 63 | 120 | 4:44 | 256 x 28 x 256 | 0.5 x 0.5 | 1.4 |
| T_1_w GRE | Sagittal | 3D | 7.8 | NA | 3 | 16 | 3:11 | 256 x 64 x 256 | 1 x 1 | 1 |
| MP2RAGE | Sagittal | 3D | 3500 | 900, 1400 | 2.67 | 9 | 8:17 | 240 x 64 x 256 | 1 x 1 | 1 |
| **3T thoracic spine^3^** | | | | | | | | | | |
| STIR | Sagittal | 2D | 8130 | 210 | 63 | 120 | 4:44 | 256 x 28 x 256 | 0.5 x 0.5 | 1.4 |
| T_1_w GRE | Sagittal | 3D | 7.8 | NA | 3 | 16 | 3:59 | 320 x 64 x 320 | 1 x 1 | 1 |
| MP2RAGE | Axial | 3D | 3500 | 900, 1400 | 3.48 | 9 | 10:48 | 264 x 282 x 256 | 0.8 x 0.8 | 4 |
| **7T brain^4^** | | | | | | | | | | |
| 0.5mm MP2RAGE | Axial | 3D | 6000 | 1000, 2900 | 5.14 | 4, 5 | 10:32 | 224 x 168 x 112 | 0.5 x 0.5 | 0.5 |
| 0.7mm MP2RAGE | Sagittal | 3D | 6000 | 800, 2700 | 3.02 | 4, 5 | 10:08 | 224 x 157 x 224 | 0.7 x 0.7 | 0.7 |
| T_2_^*^w EPI | Sagittal | 3D | 52 | NA | 23 | 10 | 3:40 | 220 x 180 x 88 | 0.5 x 0.5 | 0.5 |
| T_2_^*^w GRE | Axial | 2D | 4095 | NA | 11.4, 22.5, 33.6, 44.7, 55.8 | 90 | 11:26 | 240 x 168 x 30 | 0.5 x 0.5 | 0.5 |
| MoCo B_0_Co T_2_^*^w GRE | Axial | 3D | 74 | NA | 18, 29.5, 41.0, 52.4 | 10 | 11:50 | 240x180x32 | 0.5 x 0.5 | 0.5 |

^1^3T Skyra, Siemens, Erlangen, Germany, with a 20-channel head-neck coil when imaging the brain and spine in a single session or a 32-channel head-neck coil when imaging only the brain. ^2^Acquired before and ~10min after administration of 0.1mmol/kg gadobutrol. ^3^3T Skyra, Siemens, with a 16-channel spine-array coil. ^4^7T whole-body research system, Siemens, with a single-channel transmit, 32-channel phased array receive head coil.

TR: repetition time. TI: inversion time. TE: echo time. FA: flip angle. AT: acquisition time. FOV: field of view. AP: anterior-posterior. RL: right-left. SI: superior-inferior. PD/T_2_w: proton density/T_2_^*^ weighted. MP2RAGE: magnetization prepared 2 rapid gradient echoes. FLAIR: fluid attenuated inversion recovery. STIR: short-T_1_ inversion recovery. T_1_^*^w GRE: T_1_ weighted gradient recalled echo. T_2_^*^w EPI: T_2_^*^w echo planar imaging. T_2_^*^w GRE: T_2_^*^ weighted gradient recalled echo. MoCo B_0_Co T_2_^*^w GRE: navigator-guided motion and B_0_ corrected T_2_* weighted gradient recalled echo.

**Supplementary Table 2. Lesion and brain volume changes in individuals with and without disability progression.**

|  | Disability progression^1^ | No disability progression^1^ | B^2^ | P value^2^ |
| --- | --- | --- | --- | --- |
| New cortical lesion volume, µl ^3^ | 0, 0 (0-51) | 0, 1.8 (0-484) | -0.102 | 0.78 |
| Change in cortical lesion volume, µl^3^ | 21, 70 (-61 – 215) | 8.0, 51.5 (-24.1 – 490.2) | 0.831 | 0.007^#^ |
| % change in cortical lesion volume^3^ | 1.8, 7.2 (-6.5 – 154.5) | 2.1, 6.6 (-4.7 – 296.9) | 0.331 | 0.19 |
| Change in leukocortical lesion volume, µl^3^ | 0.44, 31.2 (-84.00 – 53.75) | 1.8, 7.7 (-39.3 – 483.1) | 0.078 | 0.72 |
| % change in leukocortical lesion volume^3^ | 0.9, 5.1 (-19.6 – 154.5) | 1.0, 9.8 (-8.8 – 363.6) | 0.288 | 0.29 |
| Change in intracortical lesion volume, µl^3^ | 0.25, 0.72 (-1.38 – 5.00) | 0, 1.5 (-1.4 – 6.3) | 0.024 | 0.95 |
| % change in intracortical lesion volume^3^ | 1.1, 11.2 (-12.5 – 17.5) | 0, 20.1 (-4.0 – 78.7) | -1.37 | 0.73 |
| Change in subpial lesion volume, µl^3^ | 24.31, 42.84 (-8.00 – 184.1) | 8.1, 31.9 (-3.9 – 135.4) | 1.059 | 0.003^#^ |
| % change in subpial lesion volume^3^ | 3.6, 13.1 (-11.4 – 33.9) | 1.8, 7.3 (-5.9 – 22.1) | 0.196 | 0.55 |
| New white matter lesion volume, µl^3^ | 0, 0 (0-498) | 0, 37 (0-675) | 0.094 | 0.79 |
| Change in white matter lesion volume, µl^3^ | 64.5, 138.3 (-2458 – 5274) | 37.0, 152.0 (-1688 – 2057) | 0.479 | 0.15 |
| % change in white matter lesion volume^3^ | 0.96, 1.91 (-8.95 – 11.0) | 0.54, 1.48 (-16.63 – 14.21) | 0.375 | 0.23 |
| Annualized atrophy rate (%)^4^ | | | | |
| Supratentorial brain | -0.02 ± 0.20 | 0.27 ± 0.16 | 0.294 | 0.38 |
| Cortex | -0.05 ± 0.32 | 0.14 ± 0.24 | 0.092 | 0.79 |
| White matter | 0.02 ± 0.23 | 0.35 ± 0.20 | 0.335 | 0.32 |
| Deep gray matter | 0.29 ± 2.03 | 0.31 ± 1.16 | 0.029 | 0.93 |

^1^Overall disability progression defined as an increase in EDSS of ≥1 for baseline EDSS <6 or ≥0.5 for baseline EDSS ≥6 or 20% increase in 25TW or 20% increase in 9HPT. ^2^Multivariate generalized linear model adjusted for age and sex. B represents the association between the variable of interest and progression of disability. ^3^Median, interquartile range (range). ^4^Mean ± standard deviation.  ^#^Significant after adjusting for false-discovery rate.

**Supplementary Table 3. Baseline MRI measures in people who transitioned from RRMS to SPMS vs people who remained RRMS.**

|  | **RRMS to SPMS (n=6)** | **RRMS (n=36)** | **B**^1^ | **P value**^1^ |
| --- | --- | --- | --- | --- |
| Age (years)^2^ | 52 ± 5 | 44 ± 11 | - | 0.08 |
| **Cortical lesions^3^** | | | | |
| Total cortical lesion number | 50, 71 (15-168) | 14, 22 (0-113) | 0.694 | 0.03^#^ |
| Total cortical lesion volume (μl) | 2183, 3673 (270-9888) | 321, 832 (0-6392) | 0.873 | 0.01^#^ |
| Leukocortical lesion number | 12, 13 (5-20) | 4, 11 (0-67) | 0.163 | 0.59 |
| Leukocortical lesion volume (μl) | 202, 394 (19-676) | 84, 232 (0-2818) | 0.001 | 0.99 |
| Intracortical lesion number | 3, 5 (0-9) | 1, 2 (0-20) | 0.417 | 0.23 |
| Intracortical lesion volume (μl) | 11, 20 (0-38) | 2, 8 (0-65) | 0.490 | 0.19 |
| Subpial lesion number | 35, 59 (5-142) | 5, 11 (0-93) | 0.770 | 0.02^#^ |
| Subpial lesion volume (μl) | 1789, 3850 (143-9660) | 119, 372 (0-5233) | 1.047 | 0.003^#^ |
| **White matter lesion volume (ml)^3^** | 11.5, 13.1 (4.4-26.5) | 5.7, 8.2 (0.9-50.8) | 0.162 | 0.64 |
| **Spinal cord lesion number^3^** | 6, 3.5 (5-10) | 4, 7 (0-13) | 0.575 | 0.18 |
| **Normalized brain volume (ml)^2^** | | | | |
| Supratentorial brain | 1,198 ± 49 | 1,265 ± 13 | -0.048 | 0.89 |
| Deep gray matter | 44.2 ± 4.0 | 48.8 ± 3.8 | 0.543 | 0.20 |
| Cortex | 596 ± 18 | 614 ± 7 | 0.343 | 0.42 |
| White matter | 542 ± 31 | 585 ± 8 | -0.228 | 0.84 |

^1^Multivariate generalized linear model adjusted for age and sex. B represents the association between the variable of interest and the transition from RRMS to SPMS. ^2^mean ± standard deviation. ^3^median, interquartile range (range). ^#^Significant after adjusting for false-discovery rate.

**Supplementary Table 4. Association between baseline MRI measures and annualized brain atrophy.**

|  | **Supratentorial brain^1^** | **Deep gray matter^1^** | **Cortex^1^** | **White matter^1^** |
| --- | --- | --- | --- | --- |
| Total cortical lesion number | 0.287 | 0.174 | 0.148 | 0.257 |
| Total cortical lesion volume | 0.297 | 0.215 | 0.188 | 0.220 |
| Leukocortical lesion number | 0.375*^,#^ | 0.132 | 0.095 | 0.470*^,#^ |
| Leukocortical lesion volume | 0.181 | 0.059 | 0.023 | 0.230 |
| Intracortical lesion number | 0.049 | -0.009 | -0.037 | 0.087 |
| Intracortical lesion volume | 0.060 | 0.016 | -0.018 | 0.069 |
| Subpial lesion number | 0.202 | 0.177 | 0.161 | 0.104 |
| Subpial lesion volume | 0.290 | 0.251 | 0.233 | 0.159 |
| White matter lesion volume | 0.320*^,#^ | 0.281 | 0.259 | 0.165 |
| Spinal cord lesion number | 0.046 | -0.035 | -0.044 | 0.133 |
| Normalized supratentorial brain volume | -0.205 | -0.104 | -0.102 | -0.218 |
| Normalized deep gray matter volume | -0.363*^,#^ | -0.150 | -0.232 | -0.209 |
| Normalized cortical volume | -0.241 | -0.272 | -0.273 | -0.067 |
| Normalized white matter volume | -0.224 | -0.077 | -0.074 | -0.278 |

Partial correlations adjusted for age and sex. ^1^Annualized atrophy rate. *: p<0.05. ^#^Significant after adjusting for false-discovery rate.
